# Integrating Multiple Imaging Modalities does not Boost Prediction of Carotid Artery Intima-Media Thickness in Midlife Adults

**DOI:** 10.1101/2022.01.31.22270191

**Authors:** Amy Isabella Sentis, Javier Rasero, Peter J. Gianaros, Timothy D. Verstynen

## Abstract

**Background:** Human neuroimaging evidence suggests that cardiovascular disease (CVD) risk may relate to functional and structural features of the brain. The present study tested whether combining functional and structural (multimodal) brain measures, derived from magnetic resonance imaging (MRI), would yield a multivariate brain biomarker that reliably predicts a subclinical marker of CVD risk, carotid-artery intima-media thickness (CA-IMT).

**Methods:** Neuroimaging, cardiovascular, and demographic data were assessed in 324 midlife and otherwise healthy adults who were free of (a) clinical CVD and (b) use of medications for chronic illness (aged 30-51 years, 49% female). We implemented a prediction stacking algorithm that combined multimodal brain imaging measures and Framingham Risk Scores (FRS) to predict CA-IMT. We included imaging measures that could be easily obtained in clinical settings: resting state functional connectivity and structural morphology measures from T1-weighted images.

**Results:** Our models reliably predicted CA-IMT using FRS, as well as for several individual MRI measures; however, none of the individual MRI measures outperformed FRS. Moreover, stacking functional and structural brain measures with FRS did not boost prediction accuracy above that of FRS alone.

**Conclusions:** Combining multimodal functional and structural brain measures through a stacking algorithm does not appear to yield a reliable brain biomarker of subclinical CVD, as reflected by CA-IMT.

## Introduction

Cardiovascular disease (CVD) encompasses many heart and vascular conditions that contribute to a primary cause of death for both men and women in the United States^1^. Atherosclerotic coronary artery disease is the most common CVD, with 50% of Americans older than 45^1,2^ and 10% Americans ages 33-45 living with some form of subclinical disease that prestages later clinical conditions^1,3^. In 2018, 13% of deaths in the United States were attributed to overt coronary artery disease^1^. Numerous complications of atherosclerotic CVD, including ischemia and myocardial infarction, contribute to the morbidity and mortality of the disease^4,5^.

CVD outcomes, such as myocardial infarction and preclinical markers of CVD risk, have recently been associated with functional and structural features of macroscopic brain systems. Longitudinal studies, for example, suggest that baseline metabolic activity in the amygdala predicts future myocardial infarction and components of the metabolic syndrome^6^, and that baseline levels of stress reactivity in the rostromedial prefrontal cortex are associated with future major adverse cardiovascular events^7^. Moreover, structural MRI measures of brain aging (composite measures of ventricle size, sulcal size and white matter hyperintensities) and regional cerebral blood flow relate to individual differences in the magnitude of blood pressure lowering induced by antihypertensive medication^8^, as well as the longitudinal progression of blood pressure over multiple years^9^. Lastly, functional activation in insular, anterior cingulate, medial prefrontal, hypothalamus and brainstem regions measured in response to mental stress and emotional stimuli has been shown to predict clinical CVD events^7^, mental stress-induced blood pressure reactivity^10^, and carotid-artery intima-media thickness (CA-IMT), a surrogate measure of preclinical atherosclerosis^11^.

At present, however, there is largely mixed evidence regarding the functional and structural brain imaging correlates of subclinical markers of CVD, particularly indexed by CA-IMT. Functional evidence shows, for example, that CA-IMT is associated with higher regional cerebral blood flow in some areas (medial frontal gyrus, putamen, and hippocampal regions, but also lower regional cerebral blood flow in other areas (lingual, inferior occipital, and superior temporal regions)^12^. Other findings indicate that CA-IMT associates with lower cerebral blood flow in gray matter and across the entire brain^13^. Separately from functional neuroimaging studies, there is structural brain imaging evidence indicating that CA-IMT is inversely associated with total brain tissue volume, as well as cortical tissue volume more specifically^14,15^. In parallel, however, other lines of evidence suggest no association between CA-IMT and total brain tissue volume or gray matter tissue volumes^13^. Lastly, some structural neuroimaging findings suggest an inverse association of CA-IMT and cortical thickness^16^, but again not all findings are consistent with the latter observations^17^. This heterogeneity in functional and structural brain imaging findings, as well as the isolated (unimodal) treatment of functional and structural brain imaging measures have created an open question as to whether the simultaneous (multimodal) modeling of functional and structural brain features would combine to predict a known marker of subclinical CVD and predictor of future clinical events; namely, CA-IMT. Moreover, whether such multimodal modeling would add to the prediction of subclinical CVD beyond established demographic, behavioral, and biological risk factors is unknown.

To elaborate, a majority of studies on the brain correlates of CVD risk, particularly CVD markers such as CA-IMT, use conventional analytical approaches that include univariate correlation and regression methods. A problematic feature of these methods is that they are not combined with out-of-sample validation testing, limiting inferences about model and sample generalizability. Moreover, these studies have historically relied on brain measures from a single neuroimaging modality, e.g., task-based or resting-state functional MRI, structural connectivity, metabolic activity via PET. Such unimodal analyses do not exploit or account for the distinct neurobiological properties of different neuroimaging modalities, that when combined may improve predictive power. Lastly, a focus thus far on the brain correlates of CVD risk has been on particular neural systems or networks, rather than all systems and networks across the entire brain. Taken together, it appears that integrating and combining whole-brain modalities into a transmodal machine learning model^18,19^ has the potential to overcome methodological limitations to improve the predictive utility and robustness of putative brain biomarkers of CVD risk to facilitate replication and generalization.

In the above regards, an effective biomarker or multimodal brain correlate of CVD risk would have the following characteristics. First, it would take into account the unique variability inherent to the different measures derived from imaging modalities (e.g., cortical thickness, cortical surface area, and tissue volumes derived by structural MRI, as well as dynamic activity measures reflecting neural networks derived by functional MRI). Second, it would rely on either standard clinical brain imaging sequences (e.g., T1 weighted anatomicals) or MRI data acquisition sequences that are amenable to clinical contexts and testing in diverse populations of people (e.g., resting state fMRI). Third, it would reliably predict CVD risk, not just associate with it (e.g., out of sample validation testing). Finally, a reliable brain correlate of CVD risk would account for additional variability above-and-beyond that already accounted for by other established risk factors for CVD. To these ends, the present study examined whether morphological and basic functional measures derived from T1-weighted and resting-state fMRI data could be combined in a multimodal machine learning analysis framework to reliably predict inter-individual variability in CA-IMT in a sample of neurologically healthy adults. For this we modified an identical multimodal machine learning approach used previously to predict “brain age”^20^ - a measure of brain aging when compared to chronological age that has been shown to correlate with numerous risk factors of CVD, including smoking and diabetes^21^. We then evaluated performance against the prediction of CA-IMT by Framingham Risk Scores^22^.

## Methods

### Participants

Neuroimaging, cardiovascular, and demographic data were collected from N=324 healthy participants (ages 30-51, 49% female) from the Pittsburgh Imaging Project (see Table 1). All participants provided informed consent. The University of Pittsburgh Human Research Protection Office granted study approval. Detailed information about the study population has been published in Gianaros et al., 2020^11^. This is the first report bearing on the multimodal prediction of CA-IMT from this sample and these results have not been published previously.

**Table 1:**
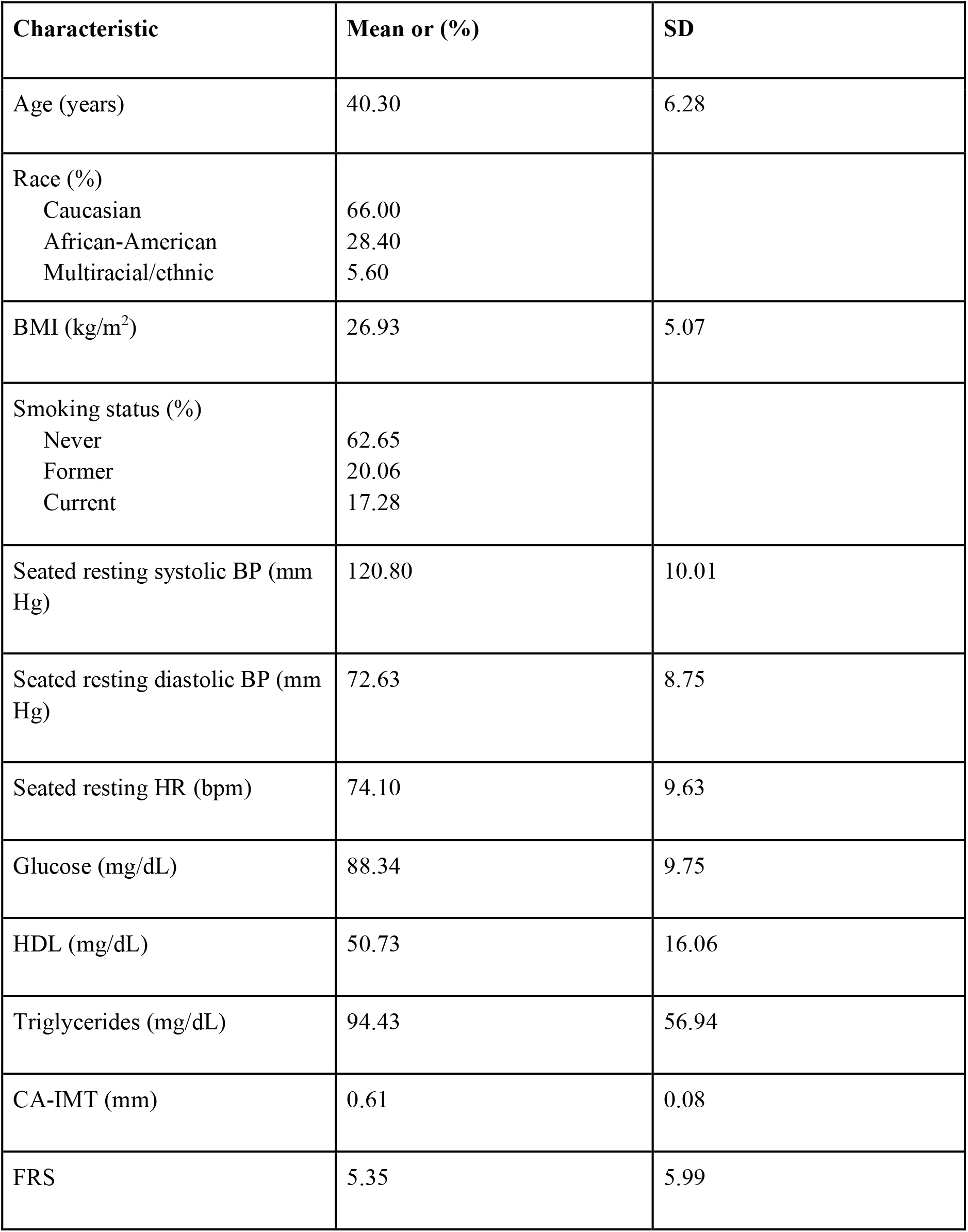
Sample characteristics (N=324; 164 Men, 160 Women). BMI = body mass index, BP = blood pressure, HDL = high-density lipoproteins, CA-IMT = carotid artery intima-media thickness, FRS = Framingham risk score.

### Preclinical atherosclerosis

Carotid artery IMT was measured at three locations (distal common carotid artery, carotid artery bulb, and internal carotid artery) by trained ultrasound sonographers using an Acuson Antares ultrasound device (Acuson-Siemens, Malvern, PA). Measurements were obtained on both the left and right carotid artery in three specific locations: 1) both the near and far walls of the distal common carotid artery, located 1 cm proximal to the carotid bulb (the location at which the near and far walls of the common carotid are no longer parallel and extending to the flow divider), 2) far wall of the carotid bulb, and 3) the first centimeter of the internal carotid measuring from the distal edge of the flow divider. These three measurements were then averaged bilaterally and across locations to calculate the mean CA-IMT, which was used as the outcome variable. Further information about measurement methods and test-retest reliability of CA-IMT measurements can be found in Gianaros et al., 2020^11^. Figure 2A shows example images of IMT acquisition. Figure 2B shows the distribution of CA-IMT values in our sample, which is approximately normal.

### Framingham risk

Framingham Risk Score (FRS) was calculated for each participant according to D’Agostino et al., 2008^22^. This metric incorporates age, sex, smoking, hypertension and cholesterol data from each participant. Five participants had missing FRS data. For analysis purposes, these missing values were imputed using the mean FRS. Figure 2C shows the distribution of FRS.

### MRI data acquisition and processing

Functional blood oxygenation level-dependent images were collected on a 3 Tesla Trio TIM whole-body scanner (Siemens), equipped with a 12-channel phased-array head coil. Resting-state functional images were acquired over a 5-minute period with eyes open and the following acquisition parameters: FOV = 205×205mm, matrix size = 64×64, TR = 2000ms, TE = 28ms, and FA = 90° (39 slices interleaved inferior-to-superior for each of 150 volumes, 3mm thickness, no gap). T1-weighted neuroanatomical magnetization prepared rapid gradient echo (MPRAGE) images were acquired over 7 min 17 sec with the following parameters: FOV = 256×208mm, matrix size = 256×208, TR = 2100ms, inversion time = 1100ms, TE = 3.31ms, and FA = 8° (192 slices, 1mm thickness, no gap).

Resting-state fMRI data were preprocessed using SPM12 and included slice-timing correction, realignment to the first image using a six-parameter rigid-body transformation, co-registration to skull-stripped and biased-corrected MPRAGE images, normalization to standard Montreal Neurological Institute (MNI) space and smoothing using a 6mm full-width-at-half-maximum (FWHM) Gaussian kernel. Head motion at the individual participant image level was estimated via framewise displacement (FD) according to Power et al., 2015^23^.

Resting-state data were denoised, including six motion parameters, white matter (WM), cerebrospinal fluid (CSF), and global signal(GS). The first principal component for each of WM, CSF and GS was used. Data were also bandpass filtered with a range of 0.009 - 0.08 Hz. A functional correlation matrix was calculated using the Craddock 200 parcellation^24^ by first computing the average time series from the voxels within each of the 200 parcels, and then calculating the z-transformed Pearson correlation coefficient between pairs of parcel time series. The upper triangular elements were extracted from the functional correlation matrix to form a vector of 19,900 functional connectivity (FC) features for each participant. FD was regressed out and the final FC vector for each participant is comprised of the resultant residuals.

MPRAGE images were analyzed using FreeSurfer (v6), with 148 cortical thickness and cortical surface area measures from the *thickness* and *area* freesurfer files respectively, using the Destrieux Atlas^25^, as well as 67 subcortical volume measures directly extracted from the *aseg.stats* freesurfer file of each participant.

### Multimodal prediction of IMT

We adopted a transmodal approach to stacking learning for prediction of CA-IMT^18,19^. In machine learning, stacking is classified as an ensemble learning method and involves combining predictions from a set of models into a new meta feature matrix for subsequent input into a new model for final prediction^20,26^.

As detailed in Figure 1, our model comprised a two-step process that used multiple output predictions for each participant from a first level support vector regression (SVR) model as the inputs into the second level random forest model. The set of first level SVR models used different groups of features, or channels, corresponding to 1) resting-state FC, 2) cortical surface area, 3) cortical thickness and 4) subcortical volume measures. Performance of the predictive models at the first and second levels of analyses was determined using cross-validation. This model was predicated on the work of Liem et al., 2016^20^, who used this transmodal approach to predict brain age. In order to validate our model implementation, we predicted brain age in our sample and compared the results to those presented in Liem et al., 2016^20^.

**Figure 1:**
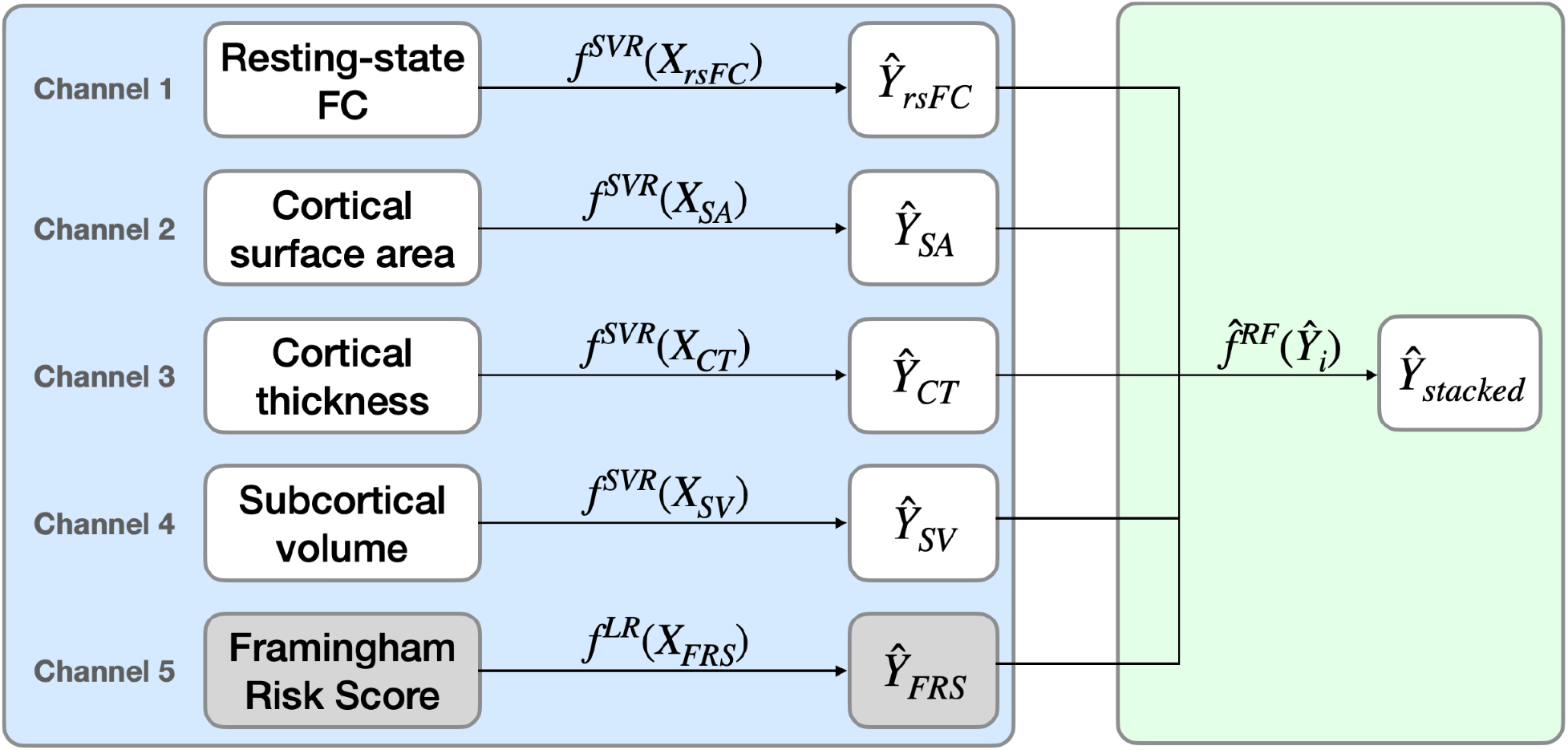
Prediction stacking model schematic, with linear SVR and linear regression used in the unimodal predictions and random forest used in the multimodal prediction. FC = functional connectivity, SVR = support vector regression, LR = linear regression, RF = random forest.

To do so, we first split the data such that 80% were used for training and the remaining 20% for testing. Next, we used five-fold cross-validation during the training stage to generate out-of-sample SVR predictions for each channel on the training set data. We used a previously tuned parameter, C, for this type of data from ^20^. As input into the second level, the out-of-sample predictions from the training set as well as the test set predictions were stacked across channels, forming new matrices of 80% observations x 4 channels and 20% observations x 4 channels, respectively. The second level random forest model was then tuned for the tree depth hyperparameter and trained using five-fold cross-validation to generate out-of-sample predictions on the new training matrix and tested on the new test matrix to generate the final predictions for brain age. Performance of the single-channel and stacked models was then evaluated by comparing participant’s chronological age with the participant’s predicted brain age in the out-of-sample test data. Prediction error was measured using the coefficient of determination, R-squared, and the root mean squared error (RMSE). All predictive analyses were performed using scikit-learn^27^.

Once validated using age, this analysis pipeline was used to predict IMT as the target outcome variable. An additional fifth channel consisting of a participant’s FRS was included in this pipeline. Since over parameterization is not a concern with a single feature model, simple linear regression (LR) was used for the single channel prediction of IMT from FRS. Thus, five single channels (four brain measures plus FRS) were stacked as input into the second level random forest model. Performance was similarly evaluated through comparison of observed IMT values with the predicted IMT values in the out-of-sample test data.

We subsequently evaluated and compared model performance on IMT prediction for every possible combination of single data channels, again using the coefficient of determination, R-squared, representing model goodness-of-fit as the measure of model performance.

Finally, in order to test robustness of our analysis and confirm that results were not dependent on a particular training/testing data split, we generated 100 random training/testing splits, using different random seeds, for analysis through our cross-validated, channel combination implementation. Final model performance was evaluated using the median of the Pearson correlation coefficient, coefficient of determination, RMSE and Bayesian information criterion (BIC) values of each partition.

## Results

We first wanted to confirm previously reported patterns in our data set. Our primary outcome measure, mean CA-IMT, was measured using ultrasound (Fig. 2A; see methods). Consistent with the assumptions of our statistical models, these CA-IMT values across our sample were approximately normally distributed (Fig. 2B), with a slight skew, in ranges consistent with an unbiased sample across the population (Stein et al., 2008). We next wanted to replicate the well established relationship between FRS and CA-IMT^28,29^. FRS values were approximately normally distributed (Fig. 2C). As expected the linear regression of the association between FRS and IMT confirms a positive association, with a Pearson correlation coefficient of r=0.3857, p<0.001. Taken together, these results are in line with the literature and constitute a replication of effects shown previously^28–30^.

**Figure 2:**
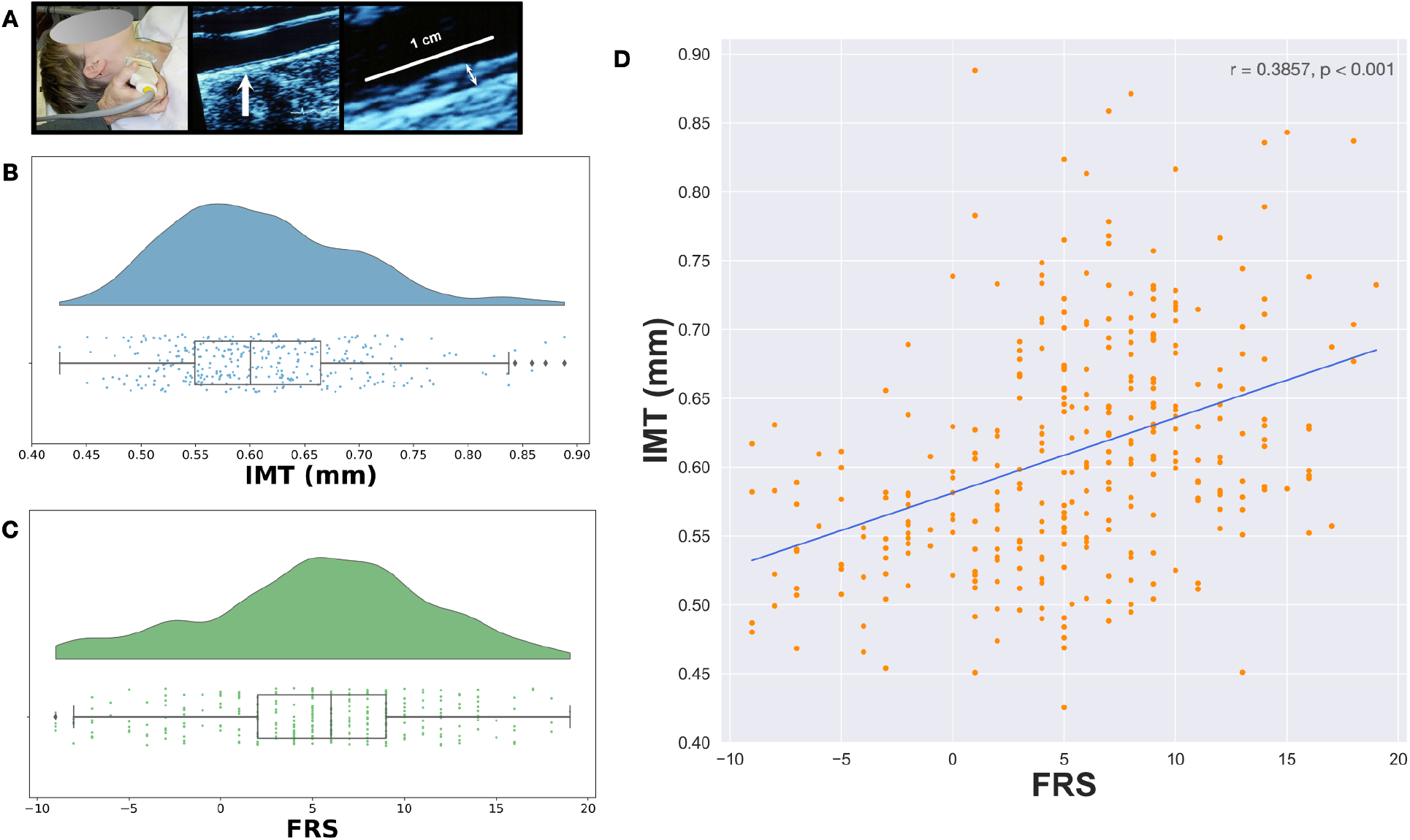
A) Left panel shows carotid artery IMT acquisition using ultrasound. Middle and right panels show example ultrasound images with the IMT indicated. B) Raincloud plot showing distribution of IMT (mm) in our sample. C) Raincloud plot showing distribution of FRS in our sample. D) Scatterplot showing the linear regression of FRS on IMT. Line of best fit shown in blue. FRS = Framingham Risk Score.

In order to validate the feasibility of our transmodal stacking approach, we first attempted to replicate the findings of Liem et al., 2016^20^ and predict chronological age using morphological brain measures as well as resting-state functional connectivity. This prior study was able to predict chronological age from the same imaging measures used here, with an accuracy of +/- 4 years. Implementing our own version of the pipeline, applied it to our sample, revealed an association between chronological age and predicted brain age that was positive and equivalent in magnitude to the original report, with a Pearson’s correlation coefficient of r = 0.5246 and coefficient of determination, R-squared = 0.2732 for the hold-out test set. Figure 3 shows the observed versus predicted scatter plot for chronological age and brain age. Our age prediction error (∼4 years) approximated that of the results presented in Liem et al., 2016^20^, confirming the validity of our stacking approach.

**Figure 3:**
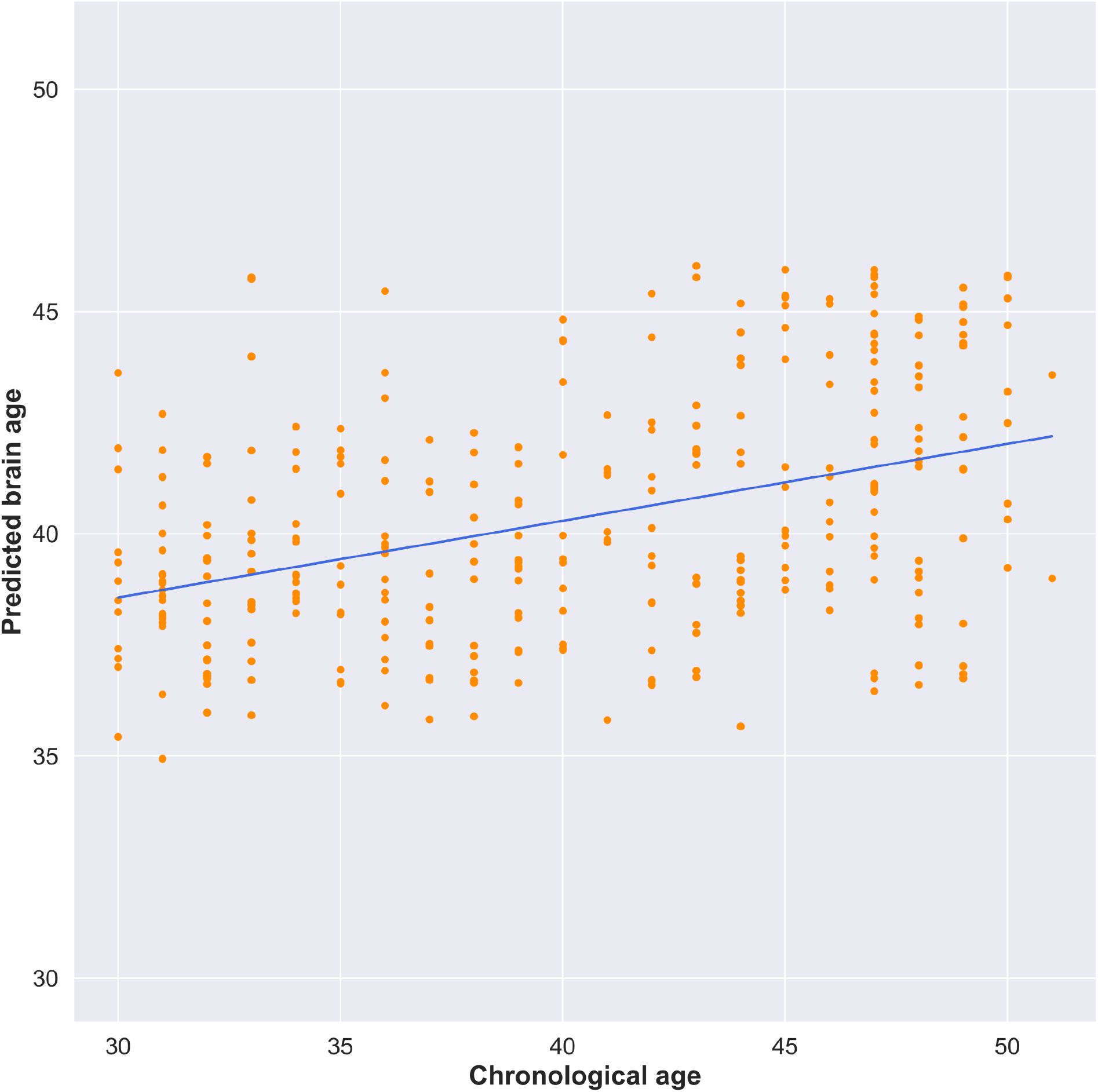
Scatterplot showing correlation between participants’ chronological age and predicted brain age according to multimodal model. Blue line represents the line of best fit.

In order to evaluate our primary aim of determining whether clinically obtainable brain imaging measures boost the prediction accuracy of individual differences in markers of CVD risk, we applied our stacked learning approach to predicting CA-IMT. Figure 4 shows the distribution of four different metrics for each random Monte Carlo data partition for both the single channel predictions of CA-IMT, as well as every possible channel combination for the second level random forest prediction of CA-IMT. Panel A shows Pearson correlation coefficients, r values, panel B shows RMSE values (with the horizontal dotted line representing the standard deviation of CA-IMT in our sample, 0.084 mm), panel C shows coefficient of determination, R-squared values, and panel D shows BIC values.

**Figure 4:**
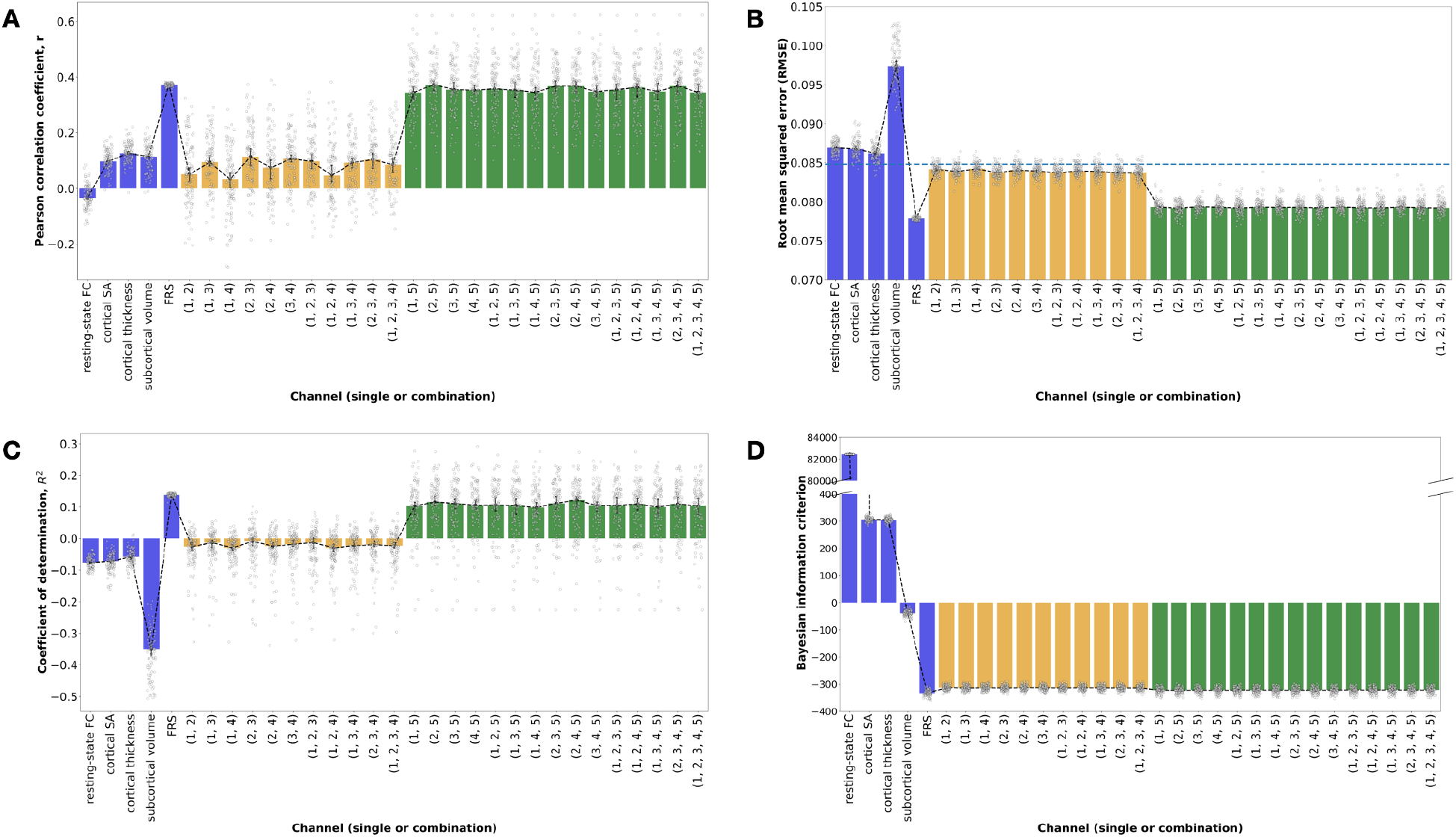
For all panels, blue bars show single channel predictions of IMT. Yellow bars show channel combination predictions that include only brain measures. Green bars show channel combinations predictions that include FRS. Error bars indicated 95% confidence intervals (calculated using 1000 bootstrap iterations). Channel combinations are indicated numerically with 1 = resting-state FC, 2 = cortical SA, 3 = cortical thickness, 4 = subcortical volume, 5 = FRS. FC = functional connectivity, SA = surface area, FRS = Framingham Risk Score. Median values for the Monte Carlo simulation for single channel and every possible channel combination prediction of mean CA-IMT: A) Pearson correlation coefficient, r, B) RMSE (horizontal dotted line represents the standard deviation of CA-IMT in our sample, 0.084 mm), C) coefficient of determination, ￼ and D) Bayesian information criterion.

Across all panels, the blue bars show the first level SVR and linear regression CA-IMT predictions using the single channel brain measures. Figure 4A shows that FRS has the largest median Pearson correlation coefficient, r = 0.3732, with the brain measures showing much smaller associations (resting-state FC median r=-0.0338, cortical SA median r=0.0970, cortical thickness median r=0.1263, subcortical volume median r=0.1145). Figure 4B demonstrates that the single channel predictions of CA-IMT from the brain measures had the largest median RMSE values (resting-state FC median RMSE=0.0869 mm, cortical SA median RMSE=0.0868 mm, cortical thickness median RMSE=0.0862 mm, subcortical volume median RMSE=0.0.0974 mm), and were higher than the standard deviation of CA-IMT (Figure 4B). The single channel FRS prediction of CA-IMT had the lowest RMSE out of all models, with a median RMSE=0.0778 mm, and was the only single channel model with an RMSE value beneath the standard deviation of CA-IMT in our sample. Figure 4C shows that median R-squared values for the single channel brain measure predictions of CA-IMT are all negative, indicating that our model does not appropriately predict CA-IMT using brain measures. However, the median R-squared values for single channel prediction of CA-IMT using FRS is positive (0.1377), indicating that FRS accounts for nearly 14% of the variance in CA-IMT. Figure 4D shows a large range of BIC values for the single channel predictions of CA-IMT, with FRS being the most negative (resting-state FC median BIC = 82,454, cortical SA median BIC = 306.18, cortical thickness median BIC = 305.06, subcortical volume median BIC = -38.99, FRS median BIC = -332.89). Note that BIC reflects the amount of information lost by a model, so lower values are better. This confirms the results from panels A-C, demonstrating that the FRS single channel model is preferred over the single channel brain measure models.

In all panels, the yellow bars show the second level random forest CA-IMT predictions from the channel combinations comprised of brain measures only. Figure 4A shows the median Pearson correlation coefficients, which ranged between r=0.0328 and r=0.1143. Stacking only the brain measures did not improve performance accuracy over the best single channel brain measure. Figure 4B shows the RMSE values, which hovered around the standard deviation of CA-IMT, and slightly improved upon the RMSE values of the single channel brain measures. Figure 4C shows that median R-squared values for the channel combination predictions of CA-IMT using only brain measures are all negative, albeit less negative that the R-squared values from the single channel brain models. This indicates that our model does not appropriately predict CA-IMT using brain measures. Figure 4D shows improved median BIC values for the channel combination predictions of CA-IMT using only brain measures compared to that of the single channel brain measure models, ranging from BIC=-312.89 to BIC=-314.22. However, these BIC values do not improve upon the median BIC value from the single channel FRS model, indicating that a combination of brain measures will not be a better feature selection choice than FRS.

In all panels of Figure 4, the green bars show the second level random forest CA-IMT predictions from the channel combinations that include FRS. Individually, some brain measures perform above chance in predicting IMT, specifically the morphometry measures from T1, when looking at the correlation between observed and predicted values. However, the effect size is smaller compared to that of the single channel FRS model. In Figure 4A, the median predicted vs. observed correlation values for the channel combinations that include FRS were more than three times that of the maximum value of the channel combinations that only include brain measures, ranging between r=0.3436 and r=0.3727. Figure 4B shows that the inclusion of FRS resulted in a reduction in median RMSE values, hovering around 0.079 mm. Figure 4C demonstrates positive median R-squared values for the channel combination predictions of CA-IMT that include FRS, ranging between R-squared=0.09 and R-squared=0.11, though all are lower than that of the single channel FRS model. Similarly, Figure 4D shows median BIC values that are smaller than that of the channel combinations that only include brain measures, but larger than that of the single channel FRS model, ranging between BIC=-320.90 and BIC=-322.23. Adding in FRS resulted in an overall increase in performance across all metrics shown in Figure 4. However, this is solely driven by FRS, as none of the channel combinations that include FRS perform better than FRS alone. These results indicate that brain measures do not assist in the prediction of CA-IMT beyond FRS.

## Discussion

Our goal for this study was to evaluate whether structural and functional brain measures from standard, clinically accessible MRI scans (T1 and resting-state fMRI) could be used to boost prediction of a marker of preclinical CVD above what is achievable from more standard clinical metrics, namely the FRS. Results show that our stacking algorithm is a sound methodology. We also see a strong association between FRS and CA-IMT, as expected. By comparison, we fail to find an improvement in our model predictions when using these brain measures individually, or in combination.

Alignment of our work with prior literature is seen in a few different ways. Firstly, we replicate existing findings that demonstrate a substantial relationship between FRS and CA-IMT^28–30^. Secondly, our methodological approach replicates that of Liem et al., 2016^20^ in a new sample, validating stacked learning as a useful tool for predicting individual differences from MRI-based measures. Finally, our results confirm some of the findings in the neuroimaging literature, namely that individually, cortical thickness and brain volumes are associated with CA-IMT^14–16^. However, these associations are weak in comparison to that of FRS and do not add to that model’s predictive power. Our findings also contrast with prior literature showing no association between CA-IMT and structural brain measures, including cortical thickness and brain volumes^13,17^, though it is possible that differences in the demographic makeup of the sample populations preclude direct comparisons.

Our failure to detect a reliable prediction of CA-IMT from the sole functional measure, resting-state FC, contrasts with recent work from our group showing reliable prediction of CA-IMT using task-based fMRI measures^11^. This contrast is particularly revealing. Resting state FC is a passive measure reflecting global intrinsic brain networks^31,32^. Thus, targeted recruitment of specific brain networks during stressful or engaging tasks is likely necessary in order to use such functional brain signals as a predictor of individual differences in CA-IMT^33^. Indeed, this type of task-based functional brain measure could boost the predictive power of FRS. However, there is a vast body of tasks that needs to be explored before this type of functional data can be incorporated into our stacking model.

An important consideration when interpreting our findings relates to our sample population. It is possible that the study selection criteria may have restricted the range of subclinical CVD present in the sample, which could partly explain the failure of multimodal brain measures to predict CA-IMT. We note, however, that FRS explained a moderate amount of the variance in CA-IMT across individuals (see Figure 4). Notwithstanding, a useful future direction would be to replicate and extend our approach in a more diverse sample, spanning a range of preclinical and clinical phenotypes of CVD.

It is also possible that predictive performance in the present study was limited by the use of CA-IMT, which has been suggested to have limited performance in the prediction of clinical CVD outcomes^34,35^. Nevertheless, evidence from intervention trials indicates that CA-IMT progression is an important outcome measure, especially for the detection of early pathophysiological vascular changes^36^. Moreover, it has been noted that carotid ultrasound is feasible in nearly all persons, relatively inexpensive, is associated with the incident (future) development of atherosclerotic plaques^37,38^. In these regards, CA-IMT is regarded as a surrogate measure of the atherosclerotic disease process that predicts later CVD events^5,37,39,40^. Taken together, while CA-IMT has advantages as a subclinical CVD marker, it is possible that predictive performance from MRI measures could be improved by using other subclinical disease markers, such as coronary calcium scores or omnibus metrics based upon CA-IMT, such as arterial stiffness and endothelial function, which reflect vascular morphology and function^41^.

The brain imaging modalities we used may have further constrained predictive performance, creating the possibility that other imaging modalities may capture brain features that are more reliably associated with subclinical CVD (e.g., arterial spin labeling for the assessment of cerebral blood flow and diffusion imaging for the assessment of white matter morphology)^42^.

In addition, our cross-sectional findings do not rule out the possibility that baseline brain measures could forecast future (prospective) changes in disease endpoints, as has been found previously. Baseline amygdalar activity has been shown to predict future occurrence of CVD events^6^, changes in visceral adipose tissue^43^ as well as risk of Takotsubo syndrome^44^. Levels of stress reactivity within the rostromedial prefrontal cortex are also associated with future adverse CVD events^7^.

In summary, the present cross-sectional human neuroimaging findings suggest that subclinical CVD reflected by CA-IMT does not reliably relate to a combined brain biomarker generated by stacking functional and structural features of the brain. Rather, CA-IMT predicted by FRS alone outperformed aggregate and individual MRI measures. In these regards, combining multimodal functional and structural brain measures by prediction stacking may not have utility in otherwise healthy midlife adults to characterize the neural correlates of subclinical CVD indexed by CA-IMT.

## Data Availability

All data produced in the present study are available upon reasonable request to the authors.

## Acknowledgments

We thank Dora Chieh-Hsin Kuan for her assistance preprocessing resting-state data and generating functional connectivity matrices. We thank Thomas Kraynak for his assistance calculating framewise displacement.

